# Tracking Persistent Symptoms in Scotland (TraPSS): A Longitudinal Prospective Cohort Study of COVID-19 Recovery After Mild Acute Infection

**DOI:** 10.1101/2024.03.07.24303931

**Authors:** Nicholas F Sculthorpe, Marie Mclaughlin, Luke Cerexhe, Eilidh Macdonald, Antonio Dello Iacono, Nilihan EM Sanal-Hayes, Joanne Ingram, Rachel Meach, David Carless, Jane Ormerod, Lawrence D Hayes

**Affiliations:** Sport and Physical Activity Research Institute, School of Health and Life Sciences, University of the West of Scotland, Glasgow, G72 0LH, UK; School of Health and Society, University of Salford, Salford, M6 6PU, UK; School of Education and Social Sciences, University of the West of Scotland, Glasgow, G72 0LH, UK; Long-COVID Scotland, 12 Kemnay Place, Aberdeen, AB15 8SG, UK; Lancaster Medical School, Lancaster University, Lancaster, Lancaster, LA1 4AT, UK

**Author notes:** Correspondence to Lancaster Medical School, Health Innovation One, Faculty of Health & Medicine, Sir John Fisher Drive, Lancaster University, Lancaster, LA1 4AT, UK.

**Keywords:** Long-COVID, post-acute sequelae of SARS-CoV-2 (PASC), tracking, symptoms, app

## Abstract

**Background:** COVID-19 disease results in disparate responses between individuals and has led to the emergence of Long-COVID, characterized by persistent and cyclical symptomology. To understand the complexity of Long-COVID, the importance of symptom surveillance and prospective longitudinal studies is evident.

**Methods:** A 9-month longitudinal prospective cohort study was conducted within Scotland (n=287), using a mobile app to determine the proportion of recovered individuals, those with persistent symptoms, common symptoms, and associations with gender and age.

**Results:** 3.1% of participants experienced symptoms at month 9, meeting the criteria for Long-COVID, as defined by the NICE terminology. Fatigue, cough, and muscle pain were the most common symptoms at baseline, with fatigue persisting the longest, while symptoms like cough improved rapidly. Older age increased the likelihood of reporting pain and cognitive impairment. Female gender increased the likelihood of headaches and post-exertional malaise (PEM), and increased recovery time from fatigue and PEM.

**Conclusions:** The majority of people fully recover from acute COVID-19, albeit often slowly. Age and gender play a role in symptom burden and recovery rates, emphasizing the need for tailored approaches to Long-COVID management. Further analysis is required to determine the characteristics of the individuals still reporting ongoing symptoms months after initial infection to identify risk factors and potential predictors for the development of Long-COVID.

## Introduction

Since its emergence in late 2019, the novel coronavirus, severe acute respiratory syndrome coronavirus 2 (SARS-CoV-2), has rapidly spread across the globe, with >760 million confirmed infections resulting in >6.9 million deaths [1]. Acute responses to infection vary widely, ranging from individuals who may be asymptomatic to those who experience severe respiratory distress or other organ damage[2]. The UK’s National Institute for Health Care and Excellence (NICE) has categorized the duration of these COVID-19 symptoms into three distinct phases: acute (<4 weeks), subacute (4–12 weeks), and chronic (>12 weeks), with the latter two intervals collectively recognized as ‘Long-COVID’. Prevalence estimates for Long-COVID vary, ranging from 13% in select community-based cohorts with laboratory-confirmed COVID-19, to upwards of 71% in hospitalized patients.

Given the speed with which the SARS-CoV-2 pandemic spread, early research focused on the symptoms and mechanisms of the acute infection, and it was only later studies that focused on the consequences and symptoms associated with Long-COVID. However, much early work was either retrospective or cross-sectional in nature or conducted in specialized units, providing little information about the natural history of the progression from acute infection to either recovery or Long-COVID[3]. More recently, prospective studies have emerged which provide some insight, although, these data are often limited. For example. Bai et al. [4] undertook a prospective study of patients recovering from a COVID-19 infection over a 3-month period. They reported that being female, active smoking and increased age were risk factors for progression to Long-COVID. They also reported nearly 70% of patients received a diagnosis of Long-COVID. However, their high prevalence rate was likely influenced by their data coming from a specific post-COVID outpatient service, with most patients having been both hospitalized and intubated during their acute infection. Consequently, it is unclear how well this kind of prospective data reflects the natural progression of the condition in many people who remained community-dwelling and were never hospitalized during the acute phase.

Other prospective studies have reported similar prevalence rates over 6- [5] or 12-months [6] using larger, more representative samples, and other meta-analyses and data pooling resulted in smaller estimates [7,8]. While these studies provide valuable estimates of prevalence, they provide limited information regarding the natural history of COVID-19 infection. Indeed, few studies have examined the longitudinal evolution of symptoms starting from the point of acute infection. Most have only a single follow-up, making it difficult to assess the longitudinal changes in symptom load. Finally, most studies only undertake symptom counting and fail to include broader assessments of patient-reported outcome measures (PROMS) such as the Long-COVID Core Outcome Set (LC-COS) using validated instruments at regular time points[9].

A clearer picture of the natural history and long-term sequelae after COVID-19 infection is needed to inform management and treatment. Therefore, the aim of this project was to track symptoms of individuals following a COVID-19 infection for 9-months using a specially designed mobile health app to determine symptomology changes over time and to undertake regular assessments with validated instruments. Our objectives were (1) to evaluate the natural history of symptoms post-infection, (2) to detect the proportion of people who have persistent symptoms, (3) to identify the most common symptoms associated with COVID-19 recovery and their relative frequencies, and (4) to identify associations of gender and age with symptom recovery.

## Methods

Ethical approval was granted from the University of the West of Scotland ethical approval committee (approval number 14019). Informed consent was provided via a mobile app signature function. Participants could withdraw from the study at any time without giving reason.

### Study design

A 9-month longitudinal prospective tracking study was conducted within Scotland using a bespoke mobile app – ‘TraPSS’. Once per month, participants were required to ‘check-in’ by completing a set of instruments within the app, which contained questions regarding COVID-19 symptoms, validated questionnaires regarding general health and well-being, and a cognitive function test. Participants were sent a notification reminder to complete the app every 31 days but were able to complete the app as often as they wished. At each check-in, responses took ∼20 minutes to complete.

### Patient involvement

Long COVID Scotland were the PPI partners for the project and became involved prior to the initial proposal. JO became part of the project team and liaised with LC Scotland’s members regarding the design, the selection of instruments, and useability testing of the mobile tracking app. As part of the project team JO advocated for participants during discussions of study management and progression. Following the project’s completion and publication of final results, further feedback will be provided to LC Scotland, who will support further dissemination of the findings through their networks.

### Inclusion criteria

Participants were included in the study if they were adults (≥18 years) living in Scotland and had a positive COVID-19 test within the previous 10 weeks. Respondents were excluded for insufficient English language for messaging to be effective; no mobile device access; impaired cognitive function which compromized comprehension of study information or messaging; current participation in any COVID-19 intervention, receiving therapies known to cause symptom exacerbations (e.g. chemotherapy).

### Participant recruitment

Participants were recruited via snowball sampling using social media posts on Facebook/Meta and Twitter/X between January 2022 and January 2023. Expressions of interest from participants were met with an invitation to a one-to-one virtual meeting with a member of the research team. Here, the study was fully explained, and the researcher determined participant eligibility to participate. This meeting also gave participants a chance to ask questions or raise concerns before participating.

### Protocol for downloading the app

The research team provided instructions and technical guidance on downloading and completing the TraPSS app on an Android or iOS device. Participants searched for the app on iOS or Android devices using the search term ‘TraPSS’. Once downloaded, participants created an account using a personal email address. Once downloaded, the app took participants through a set of onboarding screens designed to explain the features of the app. Completion of onboarding required participants to consent to take part in the study and provide a digital signature to proceed. A screengrab of the consent screen and signature was captured and remotely stored. After completing consent, the app gathered some basic demographic data, including gender, height, body mass, underlying medical conditions, and vaccination status.

### App design

At the time of development, the LC-COS had not been finalized. However, the initial Delphi survey had been completed, outlining the relevant key domains and, consequently, instruments included in the app reflected these [9]. Where we subsequently refer to mapping to LC-COS in this manuscript, we mean we mapped to the LC-COS domains. The team were also mindful to select instruments that were both valid but minimized participant burden. Consequently, the main interface was split into four sections, with each section to be completed at least once per month. Data collected via the app was stored on a GDPR-compliant server, with data accessed and downloaded using an automated Python script. In addition, each day, the server sent reminder notifications to participants who had not yet completed that week’ questions.

The four sections of the main screen comprized a symptom check-in, two sets of validated instruments grouped into ‘set A’ and ‘set B’ for ease of access, and a cognitive test. The symptom check-in included a single-item assessment mapping to LC-COS recovery (adapted from Tong et al. [10]) and single response items regarding changes in work circumstances (LC-COS work/occupational changes), and the ability to report new COVID-19 infections. Finally, participants could identify current symptoms from a list of 14 based on a scoping review of symptoms [3] and report the frequency (days/week) with which they experienced the symptom. If participants had symptoms not on the list, they could speak or type additional symptoms into the app.

Question set A included assessments of the quality of life using the SF12 [11] (LC-COS Physical functioning), the presence of post-exertional malaise (PEM) using the modified PEM Questionnaire[12] (LC-COS Post-exertion symptoms) and the Edinburgh Neurological Questionnaire (ENS) to assess for the presence of other neurological symptoms [13] (LC-COS nervous system functioning).

Question set B included the MRC dyspnoea scale [14] assessing breathlessness/dyspnoea against the ability to carry out activities of daily living (LC-COS respiratory functioning); the European Quality of Life-5 Domains (EQ-5D) [15] to assess anxiety/depression, mobility, pain, self-care, and activity (LC-COS mental functioning), the Patient Health Questionnaire-4 [16] (PhQ4) to assess anxiety and depression where scores a score ≥3 for first 2 questions suggests anxiety and a score ≥3 for last 2 questions suggests depression (LC-COS mental functioning). Set B also included a visual analogue scale to grade pain on a scale of 0 (no pain at all) to 100 (worst pain imaginable), and self-management was assessed using the Self-Efficacy for long-term conditions[17] which graded self-efficacy on a scale of 0 to 100 for disease-specific self-confidence.

Finally, the fourth section of the app contained a Single Digit Modalities Test (SDMT)[18] (LC-COS Cognitive Functioning) for a measure of cognitive function. A series of individual shapes appear on the screen, and the participant must correctly identify which number (0-9) the current shape corresponds to according to the grid at the top of the screen. Participants were scored based on the number of correctly identified shapes and the time to completion.

### Data handling

Data collected via the mobile app was stored as anonymized files on a cloud-based protected server to which only the research team had access. Python script was used to download the data from the server into CSV files, which were converted into Excel for initial data cleaning.

### Statistical analysis

Descriptive data is presented as mean ± standard deviation (SD) unless otherwise stated for demographics including gender and age, underlying health condition, and vaccination status. To examine the effects of time, age, and gender on the construct of recovery, we used the following linear mixed-effects model:

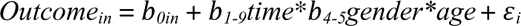

All domains of recovery represented the repeated-measures *outcome* for subject*_in_* and served as outcome measures whereas *time* (continuous variable with 9 levels [consecutive months]), *gender* (categorical variable with 2 levels [female and male]) and age (continuous variable) were modelled as predictors and treated as fixed effects alongside their three-way and two-way pairwise interactions. Moreover, random effects were assumed for *participants*, with random slopes per the predictor time introduced in the model as this addition did not result in a convergence error. We made the assumption that data were missing at random. Estimated marginal means and 95% confidence intervals were calculated alongside comparisons made using post-hoc Holm-Bonferroni adjustments. Visual inspection of residual plots was used to confirm the assumptions of homoscedasticity or normality, which was also assessed through the Shapiro-Wilk test. Moreover, since regression models can be sensitive to multicollinearity, we computed the variance inflation factors for all predictor parameters used in the linear mixed-effects model to inspect the presence of autocorrelation between pairs of predictors. Model residuals were qualitatively examined for structure and heteroscedasticity. We computed 90% CIs of the adjusted effects using the bias-corrected and accelerated bootstrap with 5,000 replicates. All statistical analyses were conducted in R language and environment for statistical computing using the *lme4*, *lmerTest*, *emmeans*, and *ggeffects* packages while model assumptions were checked using the *performance* package (4.0.5; R Core Team, Vienna, Austria). GraphPad Prism 9 was used to create all figures.

## Results

287 participants were enrolled into the study. The mean duration from initial infection to enrolment (baseline) was 35 ± 19 days.

**Table 1.** Participant Demographics.

|  | **Combined**<br>**(n = 287)** | **Males**<br>**(n = 63)** | **Females**<br>**(n = 224)** |
| --- | --- | --- | --- |
| Age (years; Mean ± SD; range) | 43.4 ± 12.4<br>(20-72) | 43.5 ± 12.4<br>(20-72) | 43.4 ± 11.9<br>(21-69) |
| Underlying health conditions (%) |  |  |  |
| Yes | 116 |  |  |
| Asthma | 41 | 7 | 34 |
| Breathing problems or other lung conditions | 1 | 1 | 0 |
| Cancer | 6 | 3 | 3 |
| Diabetes | 10 | 4 | 6 |
| Heart disease | 5 | 3 | 2 |
| Hypertension | 17 | 9 | 8 |
| Kidney disease | 1 | 0 | 1 |
| Other | 60 | 12 | 48 |
| Vaccination Status (%) |  |  |  |
| Yes - 2 doses plus booster | 234 | 52 | 182 |
| Yes - 3 doses | 61 | 9 | 52 |
| Yes - 2 doses | 276 | 54 | 218 |
| No | 3 | 1 | 2 |
| Yes - 1 dose | 284 | 62 | 222 |

### Infection recovery and change to work

The proportion of participants reporting ‘completely recovered’ increased from 31.7% at baseline to 96.9% at month 9 (**Figure 1**). The 3.1% not reporting ‘completely recovered at month 9, reports are of being ‘Mostly recovered’ (2.4%) or ‘about half recovered’ (0.7%). The random effects model revealed a significant time (month) effect for infection recovery (p<0.001, estimate = 0.07). Throughout the study, reporting of new infections was low, with 2% at month 1, 1.7% at month 2, 0.3% at month 3, 4, 6 and 7, and 1% at month 8.

**Figure 1.**
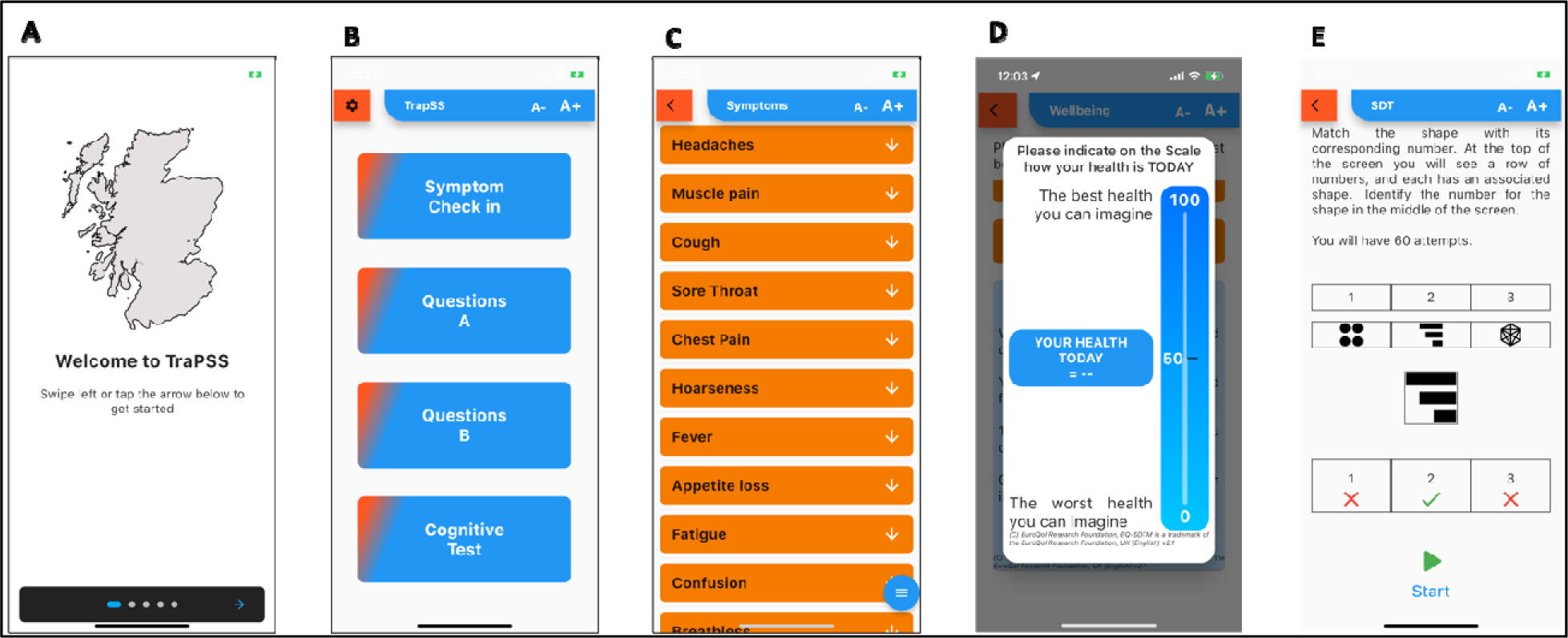
Screen shots of the bespoke Tracking Persistent Symptoms in Scotland (TraPSS) app. A – Onboarding screen, B – main home screen, C – symptom check-in screen, D – VAS scale for current health item from the Eq5D, E – instructions for the cognitive function test.

At baseline, 4.4% of participants had decreased work hours, 3.3% had stopped work completely, and 7.1% had increased work hours. Results were similar at month 9, 4.3% had decreased work hours, 3.3% had stopped work completely, and 11.6% had increased work hours. Reasons why work changed ranged from poor health (92% at baseline to 57% at month 1, 25% at month 3, 33% at month 4, 50% at month 7) to new caring responsibilities (7% at baseline to 14% at month 1, 25% at month 3) to other (0% at baseline to 28.6% at month 1, 50% at month 2, 66.7% at month 3, 100% at month 4-6, 50% at month 7). There were an insufficient number of responses to perform statistical analyses on these.

**Figure 2.**
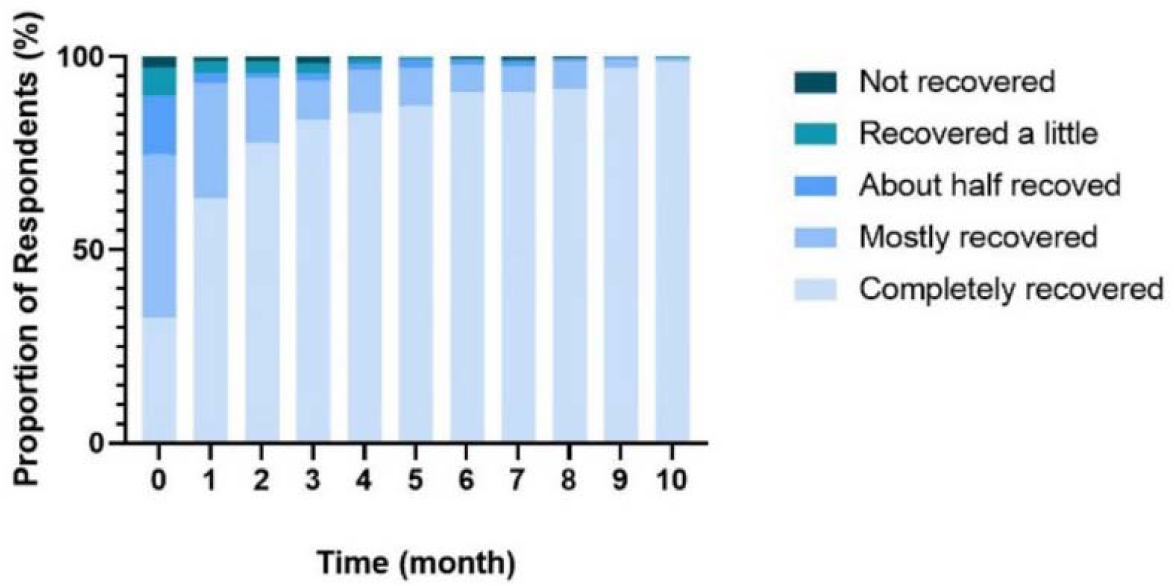
Recovery from initial infection across 9-months, based on single-item assessment (adapted from Tong et al. [10]).

### Symptom frequency

Fatigue was the most prevalent symptom at baseline, with 64.2% of participants reporting some level of fatigue. Other symptoms include headache (50.9%), cough (44.0%), muscle pain (36.9%), breathlessness (29.1%), sore throat (25.1%), confusion (23.5%), hoarseness (22.5%), chest pain (17.4%), stomach pain (14.3%), appetite loss (13.7%), fever (11.6%), loss of taste (10.9%), and loss of smell (9.9%).

At baseline, fatigue, cough, and muscle pain were the most frequently reported symptoms, with 33.8%, 25.3%, and 11.6% reporting an occurrence of 7 days/week, respectively. 64.2% of participants reported some level of fatigue at baseline. Fatigue was also the longest-lasting symptom, with 7.2% reporting some level of fatigue at month 8. Muscle pain was also long-lasting, with 5.2% of participants reporting some level of muscle pain at month 8 (decreasing from 37.9% at baseline). Cough was the fastest recovering symptom, with only 14% reporting cough frequency of 1 day/week by month 3.

The mixed-effects models revealed a significant effect of time when controlling for participants’ gender and age for most symptoms including: decreased muscle pain (p=0.004, estimate = -0.17), headache (p<0.001, estimate = -0.18), fatigue (p<0.001, estimate = -0.34), fever (p<0.001, estimate = -0.02), cough (p=0.001, estimate = -0.19), confusion (p<0.001, estimate = -0.11), breathlessness (p<0.001, estimate = -0.12), loss of smell (p<0.001, estimate = -0.05), loss of taste (p<0.001, estimate = -0.04), sore throat (p<0.001, estimate = -0.07), hoarseness (p<0.001, estimate = -0.08), chest pain (p<0.001, estimate = -0.05), stomach pain (p<0.001, estimate = -0.04), and appetite (p<0.001, estimate = -0.04). There was a significant gender [male] effect for decreased headache (p=0.024, estimate = -0.53) and fatigue (p=0.042, estimate = -0.70), and a significant gender [male] x time (month) effect for decreased fatigue only (p=0.020, estimate = 0.09). There was no significant effect of age on any symptom (all p>0.05).

**Figure 3.**
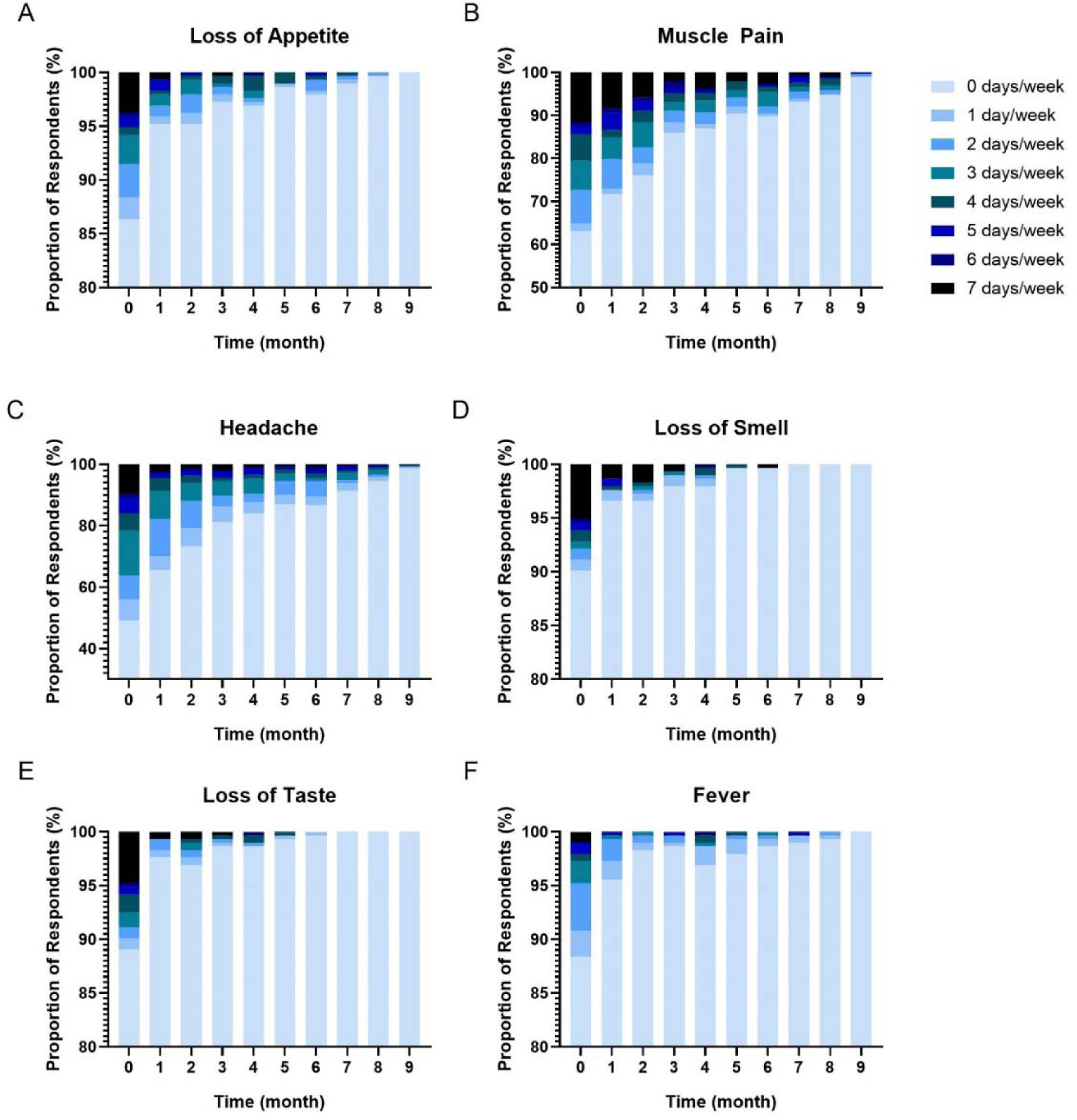
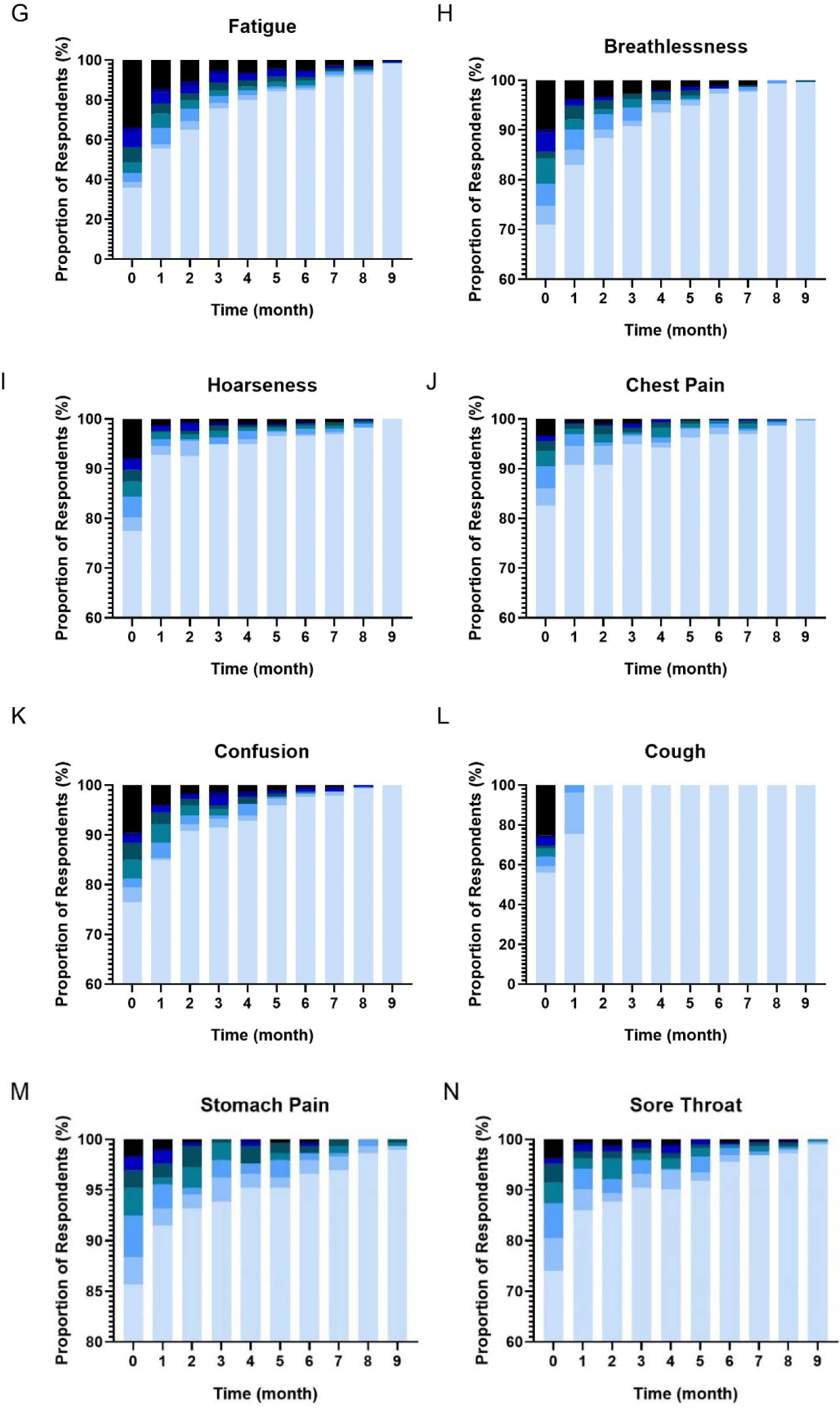
Symptom frequency over time from baseline to month 9, on a scale from 0-7 days/week, assessing loss of appetite (A), msucle pain (B), headache (C), loss of smell (D), loss of taste (E), fever (F), fatigue (G), breathlessness (H), hoarseness (I), chest pain (J), confusion (K), cough (L), stomach pain (M), and sore throat (N).

### EQ5D

From the European Quality of Life-5 Domains (EQ5D) questionnaire, 45.8% of participants reported some level of anxiety and depression at baseline, decreasing to 3.4% at month 9. Impairments in mobility and self-care are reported by 19.5% and 6.8% of participants at baseline to 1.1% and 0.7% at month 9, respectively. Some level of pain was reported by 48.5% of participants at baseline, decreasing each month to 3.1% at month 9. Activity levels had decreased in 46.4% of participants at baseline, decreasing to 1.0% at month 9. 65.6% of participants had reported less than 80% health on VAS at baseline, recovering to 2.4% at month 9. The random effects model revealed a significant time effect for EQ5D anxiety and depression (p<0.001, estimate = -0.05), mobility (p<0.001, estimate = -0.02), pain (p<0.001, estimate = -0.05), self-care (p=0.001, estimate = -0.01), activity (p<0.001, estimate = -0.05), and VAS health (p<0.001, estimate = 1.93).

**Figure 4.**
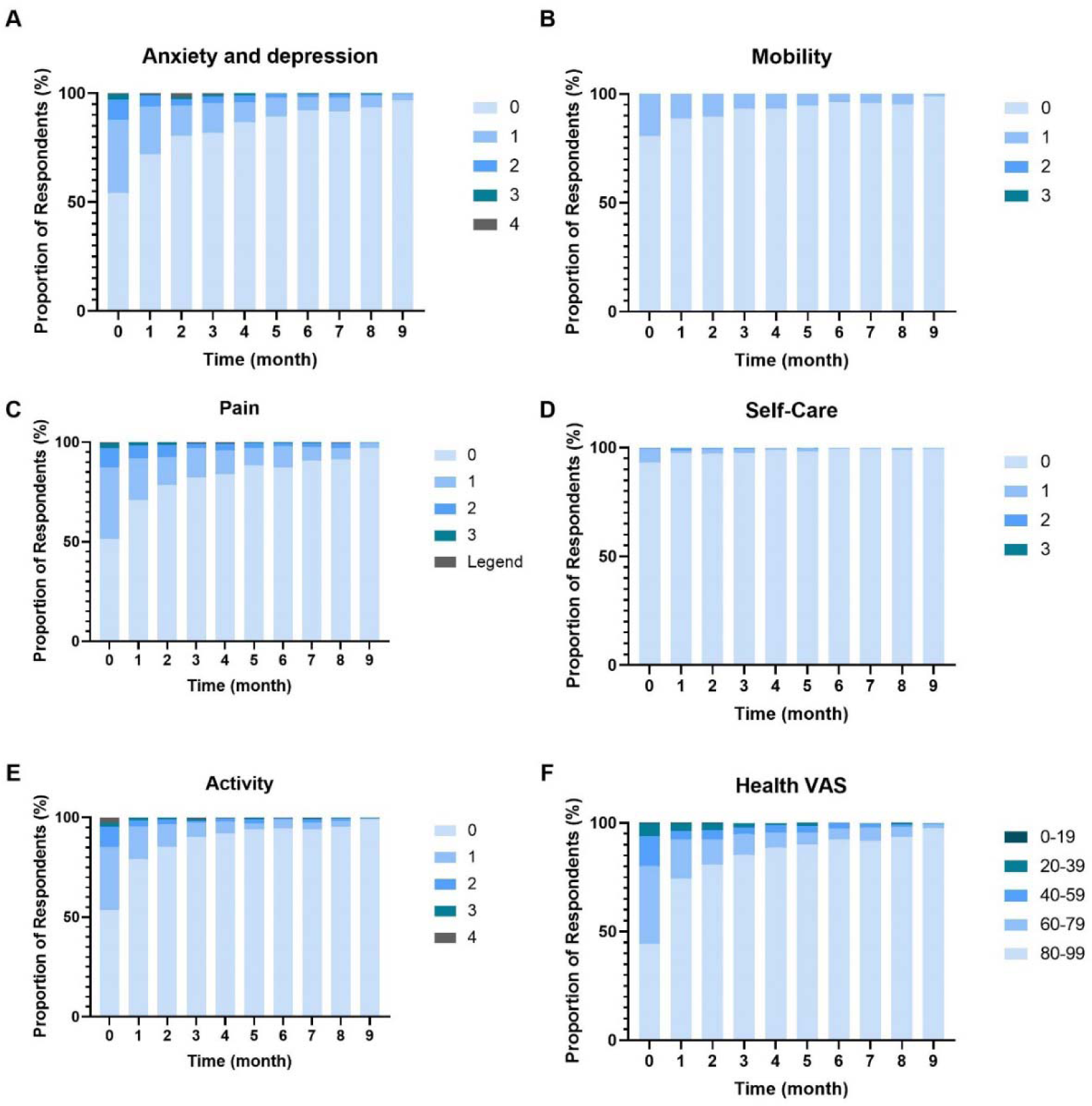
Results from EQ5D health questionnaire over time from baseline to 9-months for anxiety and depression (A), mobility (B), pain (C), self-care (D), activity (E), and health VAS (F).

### Dyspnea, VAS pain, anxiety/depression, and self-efficacy

Anxiety and depression were reported by 20.8% of participants at baseline and 0.3% at month 9. Reduced self-efficacy of condition management was experienced by 84.3% of participants at baseline and 7.2% at month 9. Dyspnea was experienced by 44% of participants at baseline, with 2.1% still reporting dyspnea at month 9. The pain was reported by 27% of participants at baseline, decreasing to 1.7% at month 9.

The random effects model revealed a significant time (month) effect for MRC dyspnea (p<0.001, estimate = -0.04), self-efficacy (p<0.001, estimate = -0.08), VAS Pain (p<0.001, estimate = -1.15), and PhQ4 anxiety and depression score (p<0.001, estimate = -1.08). There was a significant effect of age for VAS pain (p=0.028, estimate = 0.07) and PhQ4 anxiety and depression score (p<0.001, estimate = -0.88).

**Figure 5.**
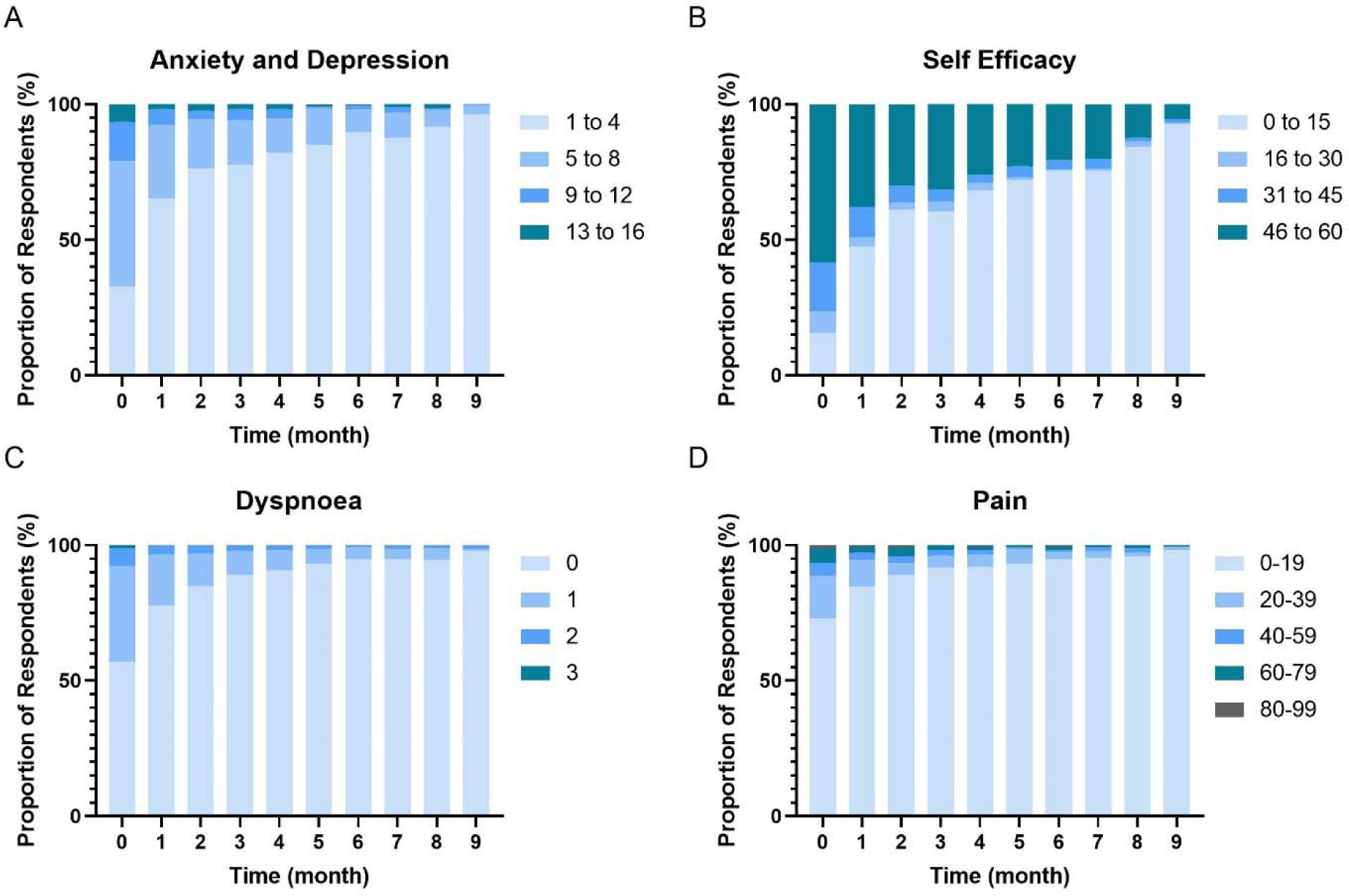
PhQ4 Anxiety and Depression (A), Self-Efficacy scores (B), MRC Dyspnea scores (C), and Visual Analogue Scale Pain (D) scores over time from baseline to 9 months.

### Post-exertional malaise

The frequency and severity of PEM decreased from baseline to month 9. At baseline, 44.0% of respondents reported mildly to severely frequent PEM. This decreased to 1.4% at month 9. The severity of PEM decreased from 19.1%, reporting mild to severe fatigue at baseline, to 0.4% at month 9. The random effects model revealed a significant time effect for PEM Severity (p<0.001, estimate = -22.64) and frequency (p<0.001, estimate = -39.16). There was a significant gender [male] effect for PEM Frequency (p=0.05, estimate = -137.68). There was a significant gender [male] x time (month) effect for PEM Frequency (p=0.033, estimate =18.96).

**Figure 6.**
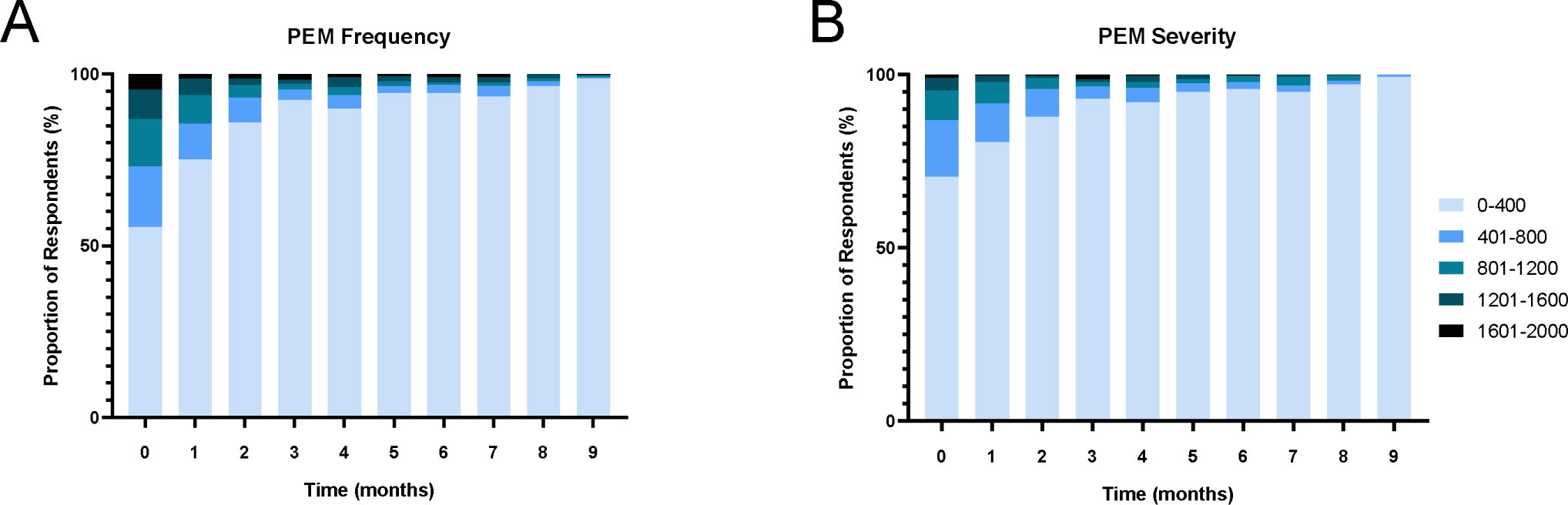
Post-exertional malaise frequency (A) and severity (B) over time from baseline to 9 months.

### Cognitive function

The number of incorrectly identified objects on the Symbol Digit Modality test reduced over time from 26.0% getting 1 or more incorrect at baseline to 1.8% at month 9. Those taking a short time (<80 s) to complete the test at baseline was 27.6% and increased to 90.4% at month 9. This was similar for total time to correct answers at 29.7% at baseline to 90.4% at month 9.

There was a significant time (month) effect for Symbol Digit Modalities total time (p<0.001, estimate = - 1.01), correct time (p<0.001, estimate = -1.08), and number incorrect (p<0.001, estimate = -0.04). There was also a significant effect of age for total time (p<0.001, estimate = 0.93) and correct time (p<0.001, estimate = 0.88).

**Figure 7.**
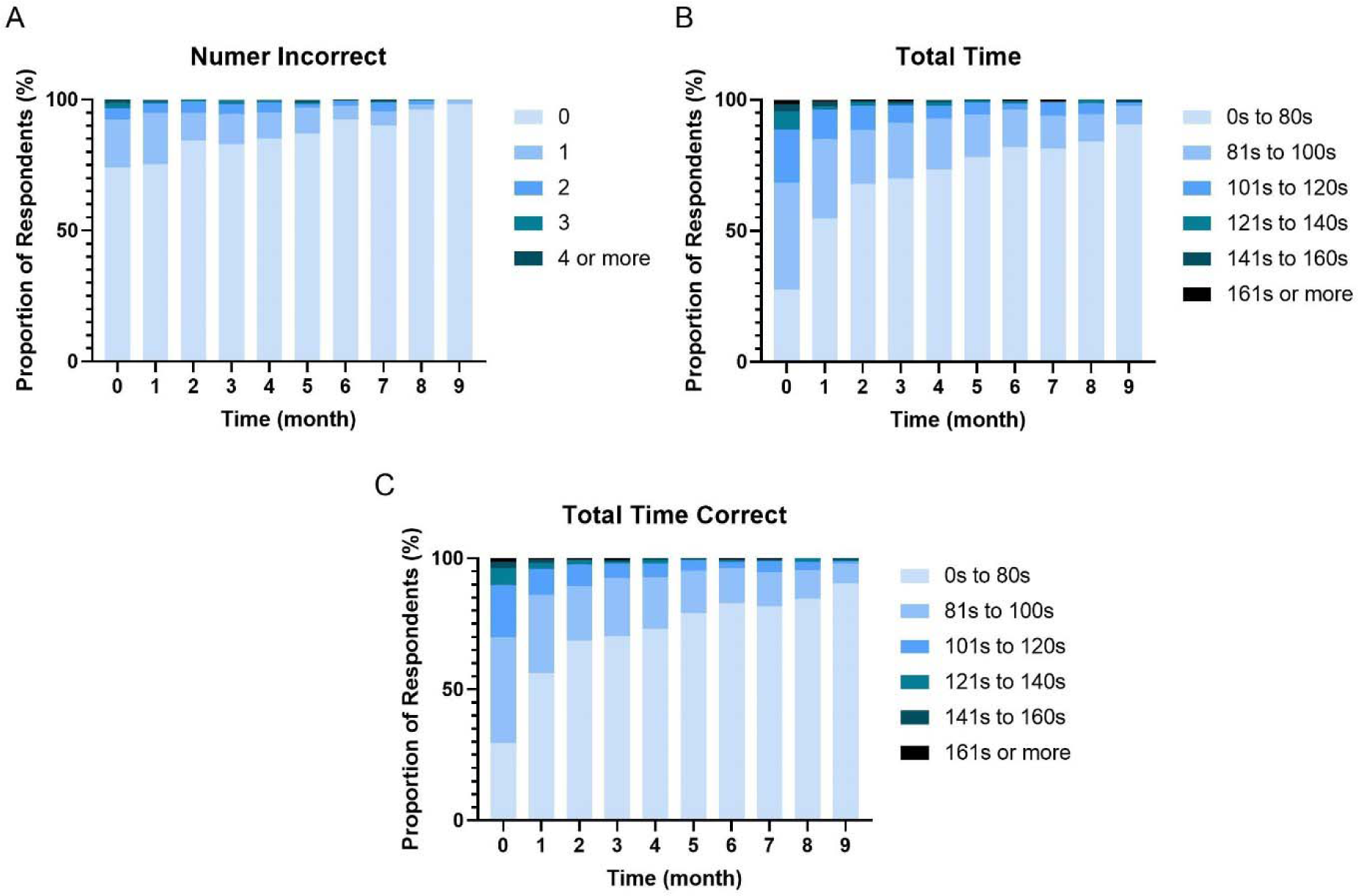
Results of Symbol Digit Modalities test from baseline to month 9, displayed as number incorrect (A), Total time to completion (B), and total time for correct answers (C) in seconds.

## Discussion

This study provides a prospective, granular view of the natural history of COVID-19, with the main findings being: (1) around a third of people self-report full recovery within the first month after infection, rising to three-quarters by the three-month meaning around a quarter of participants met the criteria for Long-COVID set out by NICE (i.e. symptoms persisting beyond 12-weeks[19]). (2) Although fatigue was the most frequent and enduring symptom, more than half of people had no symptoms or recovered relatively quickly. For some, however, recovery can be slow, with one in five still reporting symptoms after six months and one in thirty at nine months. (3) This profile, characterized by a majority experiencing minimal to no symptoms, a subset recovering slowly, and a small fraction displaying persistent symptoms throughout the study, was relatively consistent across domains. Consequently, the prevalence of long-COVID varied substantially (between 40% to 3% depending on the time frame (3 months vs 9 months post-infection) and symptoms being considered. (4) Time since infection was the only predictor of recovery for most outcomes, with males recovering more quickly for fatigue and PEM than females. Taken together, this data supports other work suggesting the recovery from COVID-19 is broadly slower than from other viral infections. Moreover, it highlights the importance of considering the duration of infection when assessing rates of long-COVID prevalence.

### Self-reported recovery and symptom load

By three months after infection, many reported partial recovery despite also reporting symptoms such as muscle pain and fatigue. This discrepancy could stem from COVID-19’s variable symptom load or individuals feeling mostly recovered except for a few persistent issues. Recovery within three months was common and higher than reported elsewhere [20,21], potentially due to our sample including only non-hospitalized individuals. Exclusively hospitalized patient studies show lower recovery rates up to a year post-infection [22,23]. By nine months, most felt fully recovered, though a small proportion (3.1%) still reported symptoms, aligning with Long-COVID criteria. This is similar to estimates of Long-COVID in Scotland from the ONS [24], and if population, it would equate to >170,000 people in Scotland with additional healthcare needs. The symptom data supports this overall view that for a sizeable minority, recovery from COVID-19 can be slow and aligns with several other studies [25,26]. Furthermore, while it is lower than some prospective studies [5,6], the differences are again likely due to those studies having a mix of hospitalized and non-hospitalized participants.

### Questionnaire responses

A novel aspect of the present study was the addition of validated psychometric instruments alongside symptom assessments. We observed distinct trajectories with all measures showing higher initial prevalence, decreasing over nine months and with a small proportion with prolonged effects.

Post-exertional malaise was reported by almost half of participants early in their recovering from COVID-19 and fell substantially over the following three months. However, it is also clear that there was a smaller cohort in whom PEM continued to occur for several months following initial infection. Most research assessing PEM has focused on individuals with Long-COVID, which explains prevalence exceeding 70% in some cohorts [27–29]. Cognitive function demonstrated a gradual improvement over 9 months, yet a subset of participants had persistent cognitive issues. Few studies have assessed cognitive function in the acute recovery phase, although cognitive dysfunction is a key marker of Long-COVID [3,8]. A recent meta-analysis did report significant reductions in executive function, attention, and working memory [30]. However, that analysis included only five studies and 290 people with Long-COVID. The present data extends this work by demonstrating that cognitive dysfunction assessed using a validated instrument, is a common feature of the acute COVID-19 response, which can persist for many months.

EQ5D assessments indicated minimal mobility or self-care limitations post-infection, though about half of participants reported experiencing anxiety, depression, and pain. Previous work has detailed more severe or similar long-term health-related quality-of-life outcomes. However, these reports have not been in cohorts of non-hospitalized participants [31,32]. Similarly, breathlessness was initially common and fell substantially within three months, with a subset experiencing persistent moderate breathlessness. Again, long-term breathlessness has rarely been studied in non-hospitalized patients. Studies in mixed or hospitalized cohorts have reported with higher [33] or similar proportions but more severe dyspnoea [34] or dyspnea. Pain assessments revealed a quarter of participants reported some degree of pain initially with prevalence falling over time. Again, comparison is difficult as previous studies have focused on hospitalized cohorts reporting higher prevalence of pain [26,35]. The present study also reported pain abated more slowly in older participants, though the effect size was small.

### Strengths and limitations

The primary strength of the present study is the use of validated instruments alongside symptom counts, at a frequency which enabled monthly tracking. This provided a granular view of recovery trajectories from acute COVID-19 infection. Moreover, we purposefully focussed on non-hospitalized individuals who have been less well represented in the COVID-19 recovery literature. There are some limitations that should also be considered. Our reliance on mobile technology and social media for recruitment may explain the lack of older participants, thus these findings may not apply to those over 70 yrs of age. While our list of symptoms was evidence based [3] and developed with our participant and patient involvement (PPI) group (Long-COVID Scotland), presenting participants with a pre-defined list of Long-COVID symptoms may have limited the range or specificity of symptoms reported by participants. Finally, reporting of new infections was low and the possibility of underreporting cannot be discounted. At that time, 1-2% of the Scottish population was testing positive for COVID-19 [36]. If underreporting was present, re-infections may have contributed to persistent or re-occurring symptoms.

### Conclusions and future directions

The main findings of this study are that around a third of individuals had no, or limited, symptoms following infection with COVID-19. Of those that did have symptoms, most recovered over the subsequent months, often much more slowly than from other viral infections. A small proportion (∼ 3%) had ongoing symptoms at the end of the 9-month follow-up. We would resist the temptation to only consider those with ongoing symptoms at the end of follow-up as having ‘true’ Long-COVID however as individuals who recovered slowly still meet the definition of Long-COVID[37] and experienced debilitating symptoms for several months after infection alongside a prolonged recovery.

What is already known:

- Infection with COVID-19 has been associated with slow recovery and persistent symptoms.
- Much of the available data is retrospective and prospective data lacks detail concerning the natural history of recovery.

What this study adds:

- This study provides evidence that for many people recovery from COVID-19 infection is slow often taking between 3-6 months.
- The slow recovery is evident across a range of domains including symptoms, quality of life, pain, and cognitive function.
- While most people do eventually recover a smaller number experience longer term persistent symptoms.

## Ethics Statement

This project received ethical approval from the Ethics Committee of the School of Health and Life Sciences at the University of the West of Scotland. Ethics number 14988

## Data availability statement

No additional data available.

## Contributors

NS, LH, JM, and JI conceived and developed the design of the study. NS developed the TraPSS app, MM, LC, EM, NS-H, RM, JO, and ADI were involved in recruitment, and data collection and analysis. NS, MM and LC drafted the manuscript. All authors reviewed and commented on the manuscript. All authors approved the final version after revision and gave permission for publication. The corresponding author attests that listed authors meet authorship criteria and that no others meeting the criteria have been omitted.

## References

1 Wong MK, Brooks DJ, Ikejezie J, et al. COVID-19 Mortality and Progress Toward Vaccinating Older Adults — World Health Organization, Worldwide, 2020–2022. MMWR Morb Mortal Wkly Rep. 2023;72:113–8.

2 Tsai P-H, Lai W-Y, Lin Y-Y, et al. Clinical manifestation and disease progression in COVID-19 infection. Journal of the Chinese Medical Association. 2021;84:3.

3 Hayes LD, Ingram J, Sculthorpe NF. More Than 100 Persistent Symptoms of SARS-CoV-2 (Long COVID): A Scoping Review. Front Med. 2021;8:750378.

4 Bai F, Tomasoni D, Falcinella C, et al. Female gender is associated with long COVID syndrome: a prospective cohort study. Clinical Microbiology and Infection. 2022;28:611.e9-611.e16.

5 Blomberg B, Mohn KG-I, Brokstad KA, et al. Long COVID in a prospective cohort of home-isolated patients. Nat Med. 2021;27:1607–13. doi: 10.1038/s41591-021-01433-3

6 Tran V-T, Porcher R, Pane I, et al. Course of post COVID-19 disease symptoms over time in the ComPaRe long COVID prospective e-cohort. Nat Commun. 2022;13:1812. doi: 10.1038/s41467-022-29513-z

7 Thompson EJ, Williams DM, Walker AJ, et al. Long COVID burden and risk factors in 10 UK longitudinal studies and electronic health records. Nat Commun. 2022;13:3528. doi: 10.1038/s41467-022-30836-0

8 O’Mahoney LL, Routen A, Gillies C, et al. The prevalence and long-term health effects of Long Covid among hospitalised and non-hospitalised populations: a systematic review and meta-analysis. eClinicalMedicine. 2023;55:101762.

9 Munblat D, Nicholson T, Williamson P. Personal Communication: Discussion of the initial outcomes of Long-COVID Core Outcome Set (COS). 2021.

10 Tong A, Baumgart A, Evangelidis N, et al. Core Outcome Measures for Trials in People With Coronavirus Disease 2019: Respiratory Failure, Multiorgan Failure, Shortness of Breath, and Recovery. Critical Care Medicine. 2021;49:503–16.

11 Huo T, Guo Y, Shenkman E, et al. Assessing the reliability of the short form 12 (SF-12) health survey in adults with mental health conditions: A report from the wellness incentive and navigation (WIN) study. Health and Quality of Life Outcomes. 2018;16. doi: 10.1186/s12955-018-0858-2

12 Cotler J, Holtzman C, Dudun C, et al. A Brief Questionnaire to Assess Post-Exertional Malaise. Diagnostics. 2018;8:66.

13 Shipston-Sharman O, Hoeritzauer I, Edwards M, et al. Screening for functional neurological disorders by questionnaire. J Psychosom Res. 2019;119:65–73.

14 Yorke J, Russell A-M, Swigris J, et al. Assessment of Dyspnea in Asthma: Validation of the Dyspnea-12. Journal of Asthma. 2011;48:602–8.

15 Vartiainen P, Mäntyselkä P, Heiskanen T, et al. Validation of EQ-5D and 15D in the assessment of health-related quality of life in chronic pain. PAIN. 2017;158:1577.

16 Löwe B, Wahl I, Rose M, et al. A 4-item measure of depression and anxiety: Validation and standardization of the Patient Health Questionnaire-4 (PHQ-4) in the general population. Journal of Affective Disorders. 2010;122:86–95.

17 Gruber-Baldini AL, Velozo C, Romero S, et al. Validation of the PROMIS® measures of self-efficacy for managing chronic conditions. Qual Life Res. 2017;26:1915–24.

18 Oirschot P van, Heerings M, Wendrich K, et al. Symbol Digit Modalities Test Variant in a Smartphone App for Persons With Multiple Sclerosis: Validation Study. JMIR mHealth and uHealth. 2020;8:e18160.

19 Overview | COVID-19 rapid guideline: managing the long-term effects of COVID-19 | Guidance | NICE. 2020. https://www.nice.org.uk/guidance/NG188 (accessed 22 August 2023)

20 Naik S, Haldar SN, Soneja M, et al. Post COVID-19 sequelae: A prospective observational study from Northern India. Drug Discoveries & Therapeutics. 2021;15:254–60.

21 Mohiuddin Chowdhury ATM, Karim MR, Ali MdA, et al. Clinical Characteristics and the Long-Term Post-recovery Manifestations of the COVID-19 Patients—A Prospective Multicenter Cross-Sectional Study. Frontiers in Medicine. 2021;8.

22 Evans RA, Leavy OC, Richardson M, et al. Clinical characteristics with inflammation profiling of long COVID and association with 1-year recovery following hospitalisation in the UK: a prospective observational study. The Lancet Respiratory Medicine. 2022;10:761–75.

23 Sigfrid L, Drake TM, Pauley E, et al. Long Covid in adults discharged from UK hospitals after Covid-19: A prospective, multicentre cohort study using the ISARIC WHO Clinical Characterisation Protocol. The Lancet Regional Health – Europe. 2021;8. doi: 10.1016/j.lanepe.2021.100186

24 Office for National Statistics. Prevalence of ongoing symptoms following coronavirus (COVID-19) infection in the UK. 2022. https://www.ons.gov.uk/peoplepopulationandcommunity/healthandsocialcare/conditionsanddiseases/bulletins/prevalenceofongoingsymptomsfollowingcoronaviruscovid19infectionintheuk/7july2022 (accessed 22 September 2022)

25 Sigfrid L, Drake TM, Pauley E, et al. Long Covid in adults discharged from UK hospitals after Covid-19: A prospective, multicentre cohort study using the ISARIC WHO Clinical Characterisation Protocol. The Lancet Regional Health - Europe. 2021;8:100186.

26 Taś A, Balo M. Post-COVID syndrome and pain perception in outpatients with COVID-19. European Review for Medical and Pharmacological Sciences. 2023;6901–10.

27 Jason LA, Dorri JA. ME/CFS and Post-Exertional Malaise among Patients with Long COVID. Neurol Int. 2022;15:1–11.

28 McLaughlin M, Cerexhe L, MacDonald E, et al. A Cross-Sectional Study of Symptom Prevalence, Frequency, Severity, and Impact of Long-COVID in Scotland: Part I - The American Journal of Medicine. 2023. https://www.amjmed.com/article/S0002-9343(23)00460-6/fulltext (accessed 24 July 2023)

29 Twomey R, DeMars J, Franklin K, et al. Chronic Fatigue and Postexertional Malaise in People Living With Long COVID: An Observational Study. Physical Therapy. 2022;102:pzac005.

30 Crivelli L, Palmer K, Calandri I, et al. Changes in cognitive functioning after COVID-19: A systematic review and meta-analysis. Alzheimer’s & Dementia. 2022;18:1047–66.

31 Kim Y, Kim S-W, Chang H-H, et al. One Year Follow-Up of COVID-19 Related Symptoms and Patient Quality of Life: A Prospective Cohort Study. Yonsei Med J. 2022;63:499–510.

32 Betschart M, Rezek S, Unger I, et al. One year follow-up of physical performance and quality of life in patients surviving COVID-19: a prospective cohort study. Swiss Med Wkly. 2021;151:w30072.

33 PHOSP-COVID Collaborative Group, Evans RA, McAuley H, et al. Physical, cognitive and mental health impacts of COVID-19 following hospitalisation – a multi-centre prospective cohort study. Infectious Diseases (except HIV/AIDS) 2021. 10.1101/2021.03.22.21254057

34 Evans RA, Leavy OC, Richardson M, et al. Clinical characteristics with inflammation profiling of long COVID and association with 1-year recovery following hospitalisation in the UK: a prospective observational study. The Lancet Respiratory Medicine. 2022;10:761–75.

35 Karaarslan F, Güneri FD, Kardeş S. Long COVID: rheumatologic/musculoskeletal symptoms in hospitalized COVID-19 survivors at 3 and 6 months. Clin Rheumatol. 2022;41:289–96.

36 Scotland PH. COVID-19 statistical report - 23 November 2022. Public Health Scotland. 10.52487/102214

37 Venkatesan P. NICE guideline on long COVID. The Lancet Respiratory Medicine. 2021;9:129.

